# *SLCO5A1* and synaptic assembly genes contribute to impulsivity in juvenile myoclonic epilepsy

**DOI:** 10.1101/2022.04.19.22273994

**Authors:** Naim Panjwani, Amy Shakeshaft, Delnaz Roshandel, Fan Lin, Amber Collingwood, Anna Hall, Katherine Keenan, Celine Deneubourg, Filippo Mirabella, Simon Topp, Jana Zarubova, Rhys H. Thomas, Inga Talvik, Marte Syvertsen, Pasquale Striano, Anna B. Smith, Kaja K. Selmer, Guido Rubboli, Alessandro Orsini, Ching Ching Ng, Rikke S. Møller, Kheng Seang Lim, Khalid Hamandi, David A. Greenberg, Joanna Gesche, Elena Gardella, Choong Yi Fong, Christoph P. Beier, Danielle M. Andrade, Heinz Jungbluth, Mark P. Richardson, Annalisa Pastore, Manolis Fanto, Deb K. Pal, Lisa J. Strug

## Abstract

Elevated impulsivity is a key component of attention-deficit hyperactivity disorder (ADHD), bipolar disorder and epilepsy^1-5^. We performed a genome-wide association, colocalization and pathway analysis of impulsivity in juvenile myoclonic epilepsy (JME). We identify genome-wide associated SNPs at 8q13.3 (p=7.5 × 10^−9^) and 10p11.21 (p=3.6 × 10^−8^). The 8q13.3 locus colocalizes with *SLCO5A1* expression quantitative trait loci in cerebral cortex (p=9.5 × 10^−3^). *SLCO5A1* codes for a membrane-bound organic anion transporter^6^ and upregulates synapse assembly/organisation genes^7^. Pathway analysis also demonstrates 9.3-fold enrichment for synaptic assembly genes (p=0.03) including *NRXN1, NLGN1* and *PTPRD*. RNAi knockdown of *Oatp30B*, the *Drosophila* homolog of *SLCO5A1*, causes both over-reactive startling behaviour (p=8.7 × 10^−3^) and increased seizure-like events (p=6.8 × 10^−7^). Polygenic risk score for ADHD correlates with impulsivity scores (p=1.60 × 10^−3^), demonstrating shared genetic contributions. *SLCO5A1* loss-of-function represents a novel impulsivity and seizure mechanism. Synaptic assembly genes may inform the aetiology of impulsivity in health and disease.

## Main

Impulsivity is a heritable behavioural trait leading to actions that are “poorly conceived, prematurely expressed, unduly risky or inappropriate to the situation and that often result in undesirable consequences”^8^. Raised impulsivity is a key endophenotype of attention-deficit hyperactive disorder (ADHD)^1^, bipolar disorder^2^ and juvenile myoclonic epilepsy^3-5^. A previous genome-wide association study (GWAS) of impulsive personality traits (UPPS-P Sensation Seeking, Drug Experimentation and UPPS-P Negative Urgency) in 22,861 healthy individuals of European ancestry demonstrated two significant associated loci at 3p12.1 and 22q13.1^9^. Variants at the 3p12.1 locus correlated with predicted *Cell Adhesion Molecule–2* (*CADM2*) expression, in the putamen^10^, and the 22q13.1 locus near *CACNA1I* has been previously implicated in schizophrenia^11^. CADM2 mediates synaptic signalling and is highly expressed in the human cerebral cortex and cerebellum^12^. Given impulsivity is elevated in neuropsychiatric disorders, there may be shared genetic architecture with impulsivity in the general population and/or across disorders, however to our knowledge there has been no GWAS of impulsivity in any neuropsychiatric disorder.

Impulsivity is elevated in different epilepsies, but the evidence across multiple dimensions of impulsivity is strongest in Juvenile Myoclonic Epilepsy (JME)^3-5^, a common adolescent-onset syndrome characterized by awakening, generalized myoclonic, tonic-clonic and absence seizures often triggered by sleep deprivation. Trait impulsivity in JME is associated with the frequency of both myoclonic and absence seizures^3^, but it is not clear if this indicates a causal relationship or a common mechanism regulating both impulsivity and seizures, though convergent lines of evidence suggest the involvement of overlapping prefrontal-striatal networks in both JME and impulsivity^13-20^. Finding a shared aetiology would offer new therapeutic approaches for drug-resistant epilepsy.

The overall syndrome of JME has complex inheritance with few replicated susceptibility loci^21^, and other loci with less support^22-24^. A major challenge for epilepsies of complex inheritance is to explain the wide variation in phenotypic expression and treatment response between individuals. Forty-percent experience antiseizure medication (ASM) resistance or intolerance^25^. In addition, no current ASM modifies the lifelong disease course of JME and the pharmacological options are sparse, especially for women^25^. Hence novel treatments based on genetic disease mechanisms, such as those emerging for monogenic channelopathy and mTOR pathway epilepsies, are urgently needed^26,27^. Our methodological approach is to carry out genome-wide analysis of endophenotypes in JME such as impulsivity and clinically relevant outcomes such as ASM resistance, a strategy with predicted advantages for reducing heterogeneity, increasing statistical power^28,29^ and improving direct clinical translation for precision medicine.

We investigated the influence of 8,950,360 variants on impulsivity in European ancestry JME patients (n=324) and a mega-analysis with all ancestries (n=372), who self-rated their trait impulsivity using the Barratt Impulsivity Scale, eight-item BIS-Brief version^30^. We conducted a GWAS of BIS-Brief score in the European subset, adjusted for sex, genotyping batch, age at consent, population stratification, and seizure frequency (Extended Data Table 1). We discovered two genome-wide significant loci at chromosome 8 (rs73293634 (G/T)) and chromosome 10 (rs75042057 (T/G) (Fig. 1 and Table 1). Association of these two loci in a mega-analysis including all ancestry groups (Supplementary Fig. 1) provides stronger evidence of association as measured by the p-value (Table 1). The phenotypic variation explained (PVE) for rs73293634 was 10.1% in the European analysis. rs73293634 falls in an intergenic region near *SLCO5A1* and two individuals with large structural deletions that include *SLCO5A1* are reported in the Decipher Genomics database with seizures and a neurodevelopmental disorder (https://www.deciphergenomics.org/gene/SLCO5A1/patient-overlap/cnvs).

**Table 1:** Summary of genome-wide associated variants and chr18 region (rs515846) for the GWAS of BIS scores in JME (n=324). All observed sample allele frequencies are comparable to those seen in the European 1000 Genomes (phase 3)^60^.

| Variant ID (hg38) | European GWAS (n=324) |  |  |  |  | Mega-GWAS (n=372) |  |  |  |
| --- | --- | --- | --- | --- | --- | --- | --- | --- | --- |
| | Imputation<br>$r^2$ | MAF | Beta | Standard<br>error | P-value | MAF | Beta | Standard<br>error | P-value |
| chr8:69,884,968*<br>rs73293634 (G/T) | 0.961 | 0.036 (T) | 5.42 | 0.91 | $7.5 \times 10^{-9}$ | 0.041 | 4.55 | 0.79 | $1.61 \times 10^{-8}$ |
| chr8:69,876,965<br>rs146866040 (A/G) | 0.979 | 0.032 (G) | 5.38 | 0.94 | $2.5 \times 10^{-8}$ | 0.031 | 5.60 | 0.90 | $1.57 \times 10^{-9}$ |
| chr10:34,202,650<br>rs75042057 (T/G) | 0.878 | 0.019 (G) | 7.51 | 1.33 | $3.6 \times 10^{-8}$ | 0.022 | 6.60 | 1.19 | $4.99 \times 10^{-8}$ |

\*The lead SNP for the mega-GWAS was rs146866040. The LD between them is 0.89.

**Fig. 1:**
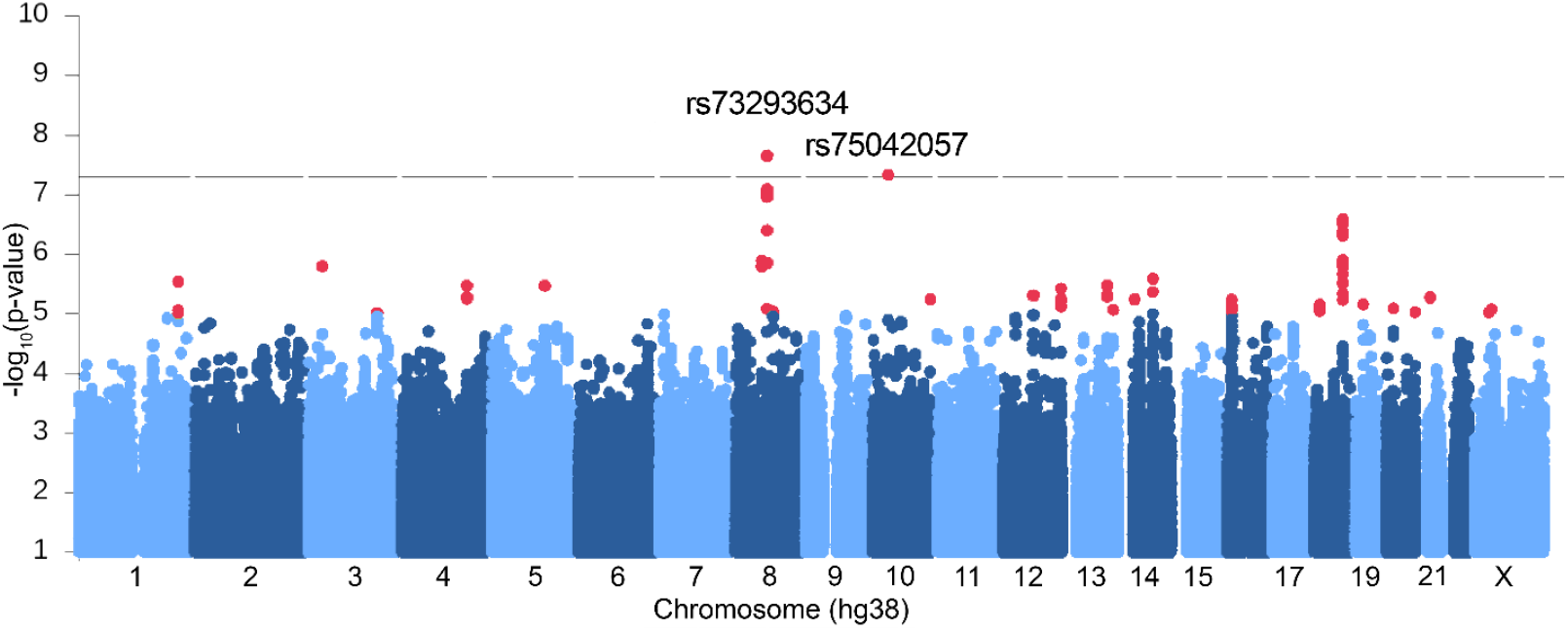
Manhattan plot showing GWAS with BIS-Brief score. We found two significant genome-wide associations on chromosome 8 (rs73293634 (G/T)) and 10 (rs75042057 (T/G)) in the analysis of 324 European individuals with JME. Variants below -log_10_P < 1 were omitted in the plot.

The significant genome-wide association on chromosome 10 (rs75042057) falls in intron 22 of *PARD3* (NM_001184785.2). The PVE by the SNP is 9.3%. Significant linkage (multipoint max LOD 4.23, alpha 0.34) was previously reported to this locus in French-Canadian families with idiopathic generalized epilepsy (IGE)^31^, of which JME is a subtype.

Since the GWAS-associated variants are not exonic, we next assessed whether the variants impact gene expression, and for which gene in which tissue of origin, by assessing colocalization of the genome-wide significant peaks with expression quantitative trait loci (eQTL) in brain tissues. We used eQTLs from the Genotype-Tissue Expression project (GTEx) v8^12^, PsychENCODE^32^, and human fetal brains^33^ and combined them with the GWAS summary statistics from the mega-analysis, for colocalization analysis adjusting for multiple hypothesis testing^34^. Colocalization analysis with eQTLs from GTEx brain and tibial nerve tissues for genes at the locus (chr8:69,650,000-70,000,000, hg38) shows significant colocalization with *SLCO5A1* in the cerebral cortex, and no colocalization with other genes in the region (Fig. 2a and Extended Data Fig. 1; Simple Sum colocalization p=9.5 × 10^−3^). The minor allele for the lead SNP rs73293634 (T) decreases expression in GTEx cerebral cortex (Fig. 2c). We found no significant colocalization in eQTLs from PsychENCODE^32^ and fetal brains^33^, although nearby variants in the locus in adult brains in PsychENCODE have, in general, a clear influence on *SLCO5A1* expression (Fig. 2b). According to BrainSpan^35,36^, *SLCO5A1* is highly expressed prenatally, with expression dropping after birth but remains detectable throughout adulthood (Fig. 2d). We did not observe significant colocalization at the chromosome 10 locus with eQTLs from adult brains in GTEx^12^, PsychENCODE^32^ or fetal brains^33^.

**Fig. 2:**
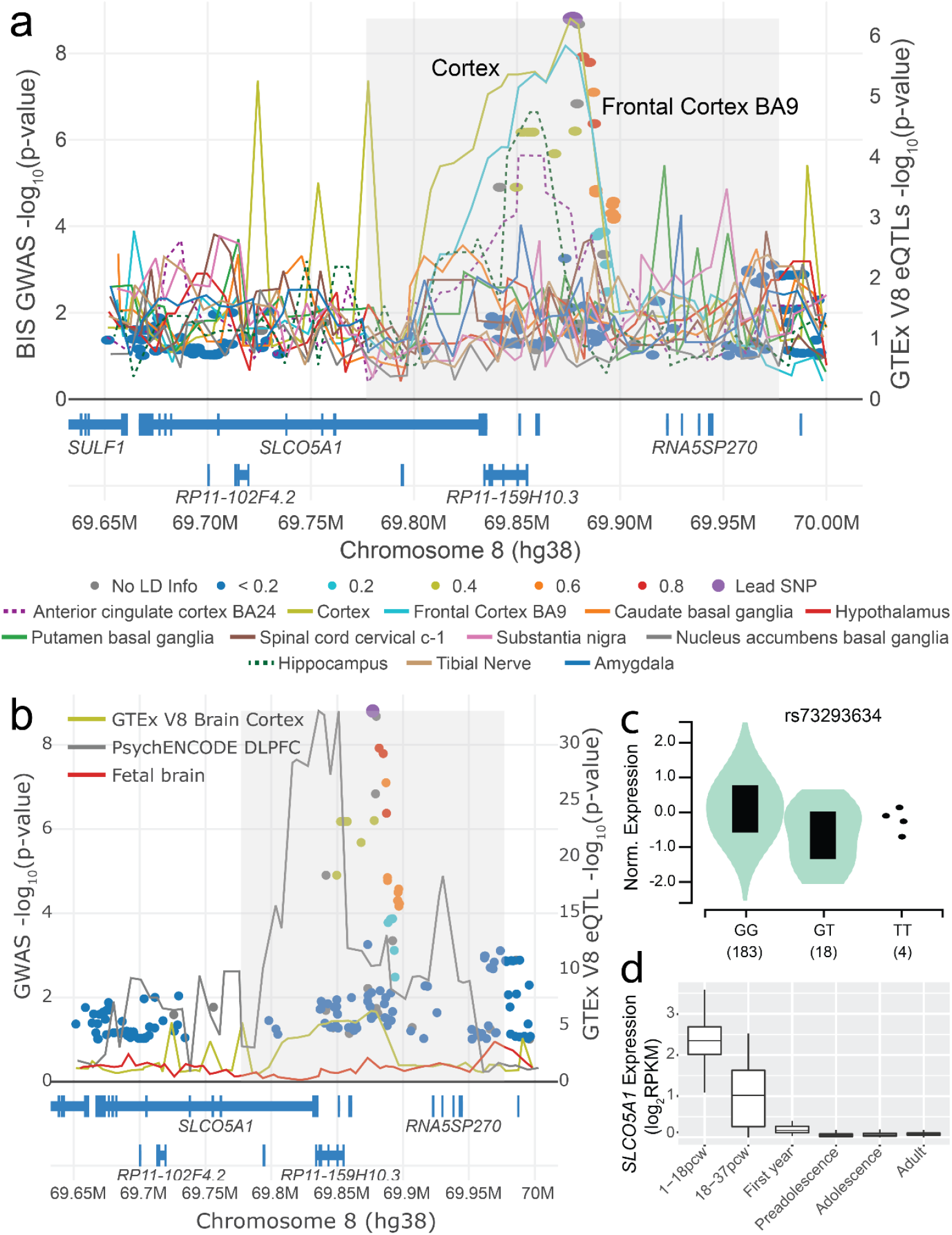
LocusFocus^65^ plot for the GWAS with BIS (dots) and eQTLs in GTEx^12^ brain and tibial nerve tissues for the *SLCO5A1* gene (lines). **a**, Colocalization figure from LocusFocus for the *SLCO5A1* gene. Lines depict the minimum p-value trace in a sliding window for *SLCO5A1* eQTLs from GTEx, one line per tissue. Circles depict the GWAS with BIS, with the lead SNP in purple and pairwise LD with the lead SNP marked as shown in the legend, calculated using the 1000 Genomes Project^60^ European subset. Significant colocalization is observed for *SLCO5A1* eQTLs in GTEx v8 for the cerebral cortex after increasing sample size in a mega-GWAS (n=367, -log_10_ Simple Sum 2^34^ P-value = 9.5 × 10^−3^). Colocalization analysis with only the Europeans is provided in Extended Data Fig. 1. Colocalization was also tested for all other nearby genes shown in the figure, but no other genes’ eQTLs colocalized with BIS GWAS (not shown). **b**, Colocalization analysis with PsychENCODE eQTLs in the dorsolateral prefrontal cortex (DLPFC) (n=1,866)^32^, and eQTLs derived from second trimester fetal brains (n=120)^33^, with GTEx’s brain cortex eQTL as in A provided for reference. Colocalization analysis results suggest no colocalization with either PsychENCODE (Simple Sum 2 P-value = 0.985) or fetal brain eQTLs (does not pass first stage test in Simple Sum 2 for having significant eQTLs in the region). **c**, Violin plot for the eQTL effect of rs73293634 SNP on *SLCO5A1* expression in the cerebral cortex from GTEx v8. **d**, Expression change of *SLCO5A1* from brains in various developmental stages from BrainSpan^35,36^. pcw, post conception weeks; preadolescence, 2-12 years old (inclusive); adolescence, 13-19 years old; adult, ≥ 20 years old (oldest samples are 40 years old).

*SLCO5A1* is a membrane-bound organic anion transporter with no known substrate^7^ (Fig. 3). We performed a full BLASTp search of the SLCO5A1 polypeptide sequence (NP_112220.2) on *Drosophila melanogaster* to identify the closest matching sequence alignment. This revealed *Oatp30B* as the closest matching gene with a 37.66% identity and E-value of 2 × 10^−150^ (NP_995667.1). BLASTp of Oatp30B polypeptide sequence (Q9VLB3) across all species for conserved domains reveals this gene has conserved Major Facilitator Superfamily (MFS), OATP, and Kazal domains (Fig. 3 and 4a). We therefore used an effective RNAi transgenic line (Extended Data Fig. 2a) to assess the effect of pan-neuronal adult knockdown of *Oatp30B/SLCO5A1*. Flies with reduced *Oatp30B* levels displayed a small but significant shortening of their lifespan (Extended Data Fig. 2b) and a striking over-reaction to vibration stimuli applied through the automated Drosophila Arousal Tracking (DART) system^37^, which elicit an otherwise modest activity response in two separate control fly genotypes (Fig. 4b). Additional analysis of locomotor behaviour clarifies that *Oatp30B* knockdown did not alter the speed of flies or the duration of each activity bout or the interval in between bouts of action (Extended Data Fig. 2c-e), indicating a specific defect in excessive response to stimuli. Furthermore, *Oatp30B* knockdown led to a dramatic increase in the frequency of seizure-like events (Fig. 4c) when exposed to hyperthermia, a trigger for seizures in *Drosophila*^38^. Recovery to full motility after seizure-like events was also significantly slower in flies with *Oatp30B* knockdown (Fig. 4d). These data establish a common causal link between *Oatp30B/SLCO5A1* downregulation, impulsivity-like startling behaviour, and susceptibility to seizure-like events.

**Fig. 3:**
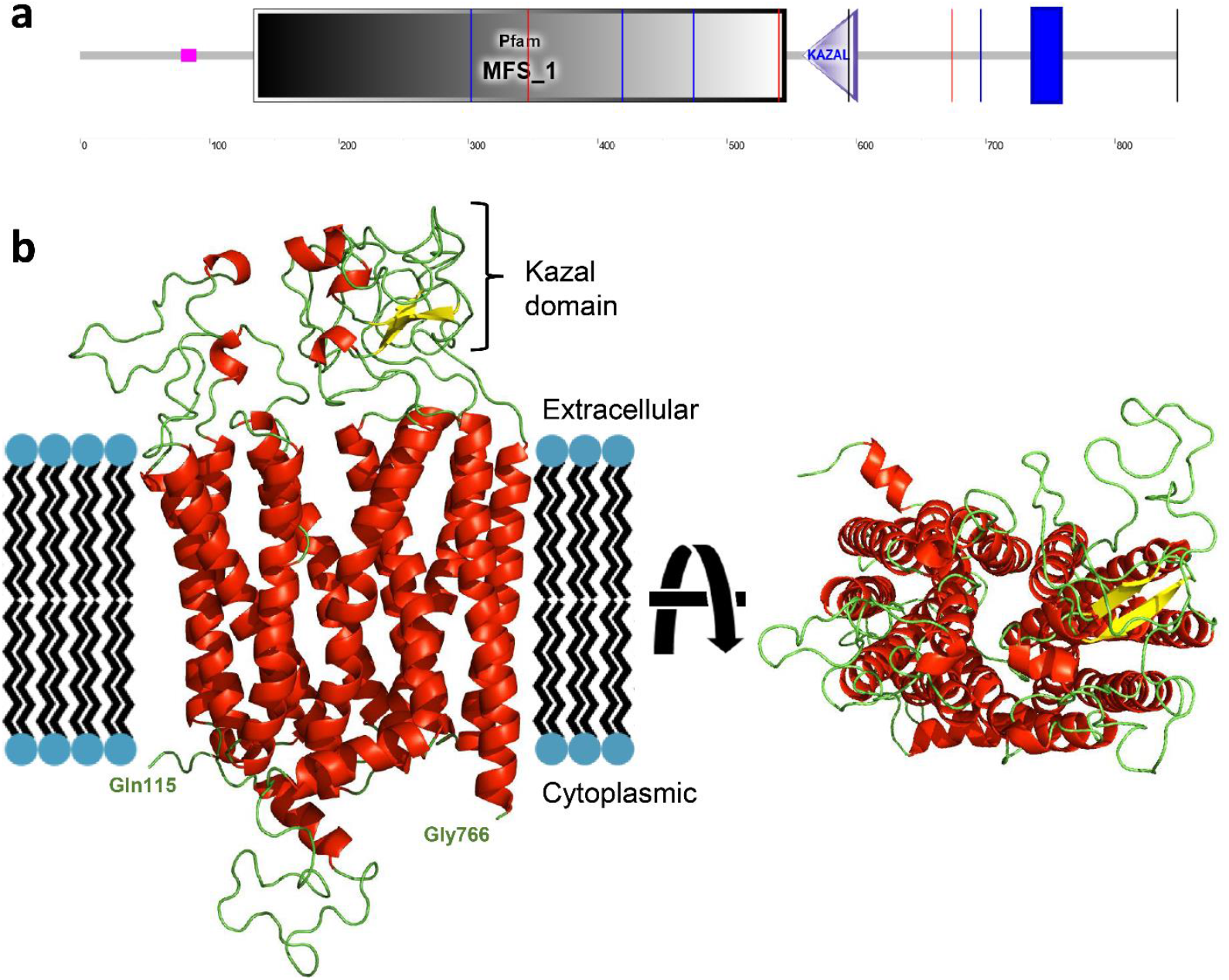
Domain architecture of human SLCO5A1. **a**, Schematic representation of the protein with the indication of recognised domains. A SMART analysis to identify structural domains confirmed the presence of two modules, Major Facilitator Superfamily (MFS) and a Kazal domain, interspaced with potentially unstructured sequences. The MFS transporters are membrane proteins capable of transporting small solutes in response to chemiosmotic ion gradients^67,68^. They are represented in many organisms from Archaea to Homo sapiens. MFS proteins target a wide range of substrates, including ions, carbohydrates, lipids, amino acids and peptides, nucleosides and other small molecules and transport them in both directions across the membrane^69^. The Kazal domain is an evolutionary conserved module usually acting as a serine-protease inhibitor. **b**, Predicted model of the monomeric form of SLCO5A1 from amino acids 115-766, built using the SwissModel homology server (https://swissmodel.expasy.org) and utilising the template structure pdb:7eeb. Red: alpha helices; Yellow: Beta strands; Green: Loops.

**Fig. 4:**
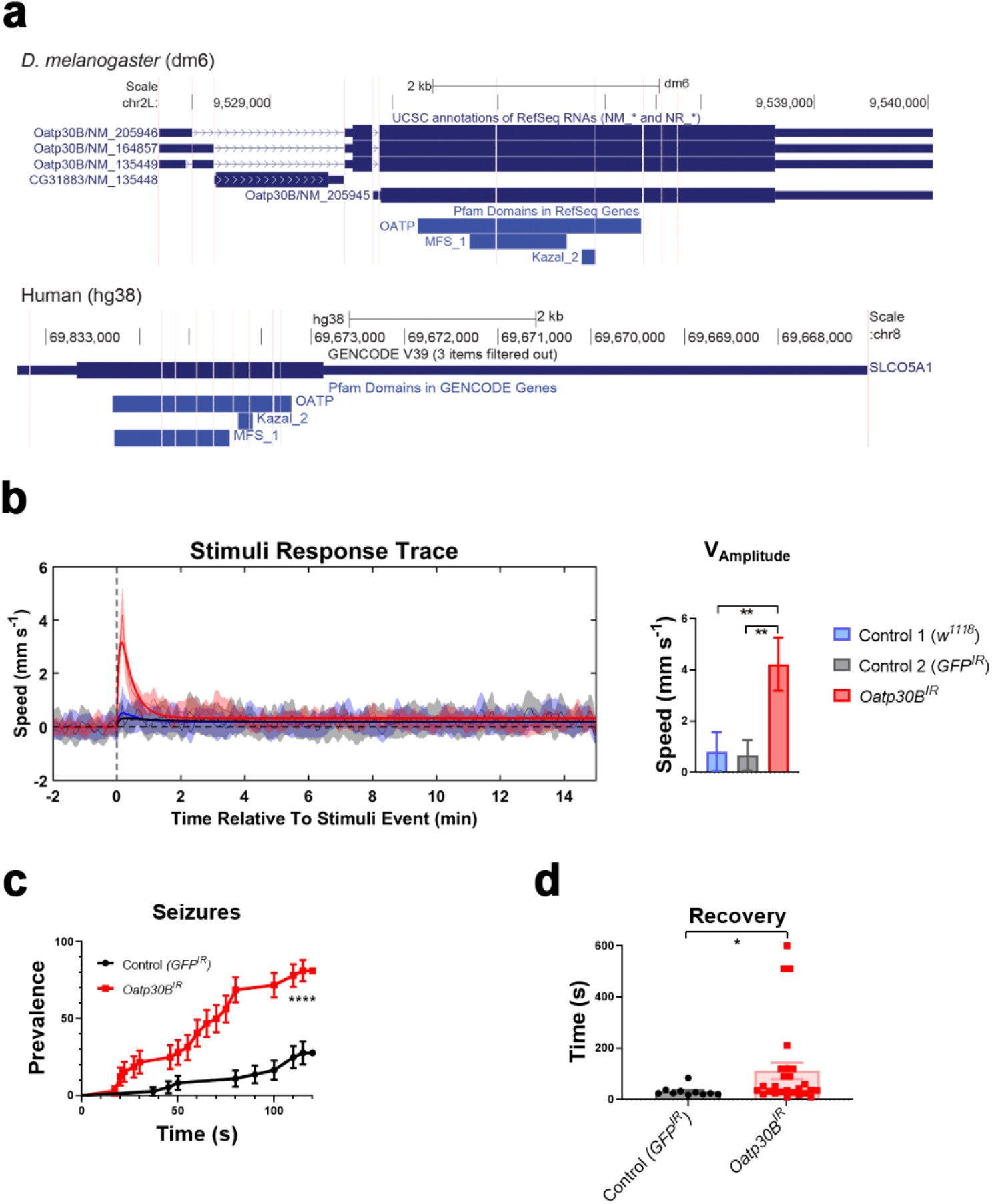
a, UCSC browser exon view of *Oatp30B* in *D. melanogaster* (dm6 assembly) and *SLCO5A1* in human (hg38 assembly). Pfam tracks show Oatp30B and SLCO5A1 have conserved MFS, OATP and Kazal domains. **b, Startling reaction to trains of vibrations in flies with *Oatp30B* knock down**. The *UAS*-*Oatp30B*^*IR*^ (GD12775) transgenic or the control *UAS-GFP*^*IR*^ were driven with *nSyb-Gal4* and *Ubi-Gal80ts*. The *w*^*1118*^ strain is a control for the genetic background in absence of transgenes. Mean +/-SEM ** p<0.01, One Way ANOVA, Tukey’s post-hoc test. Units are the vibration events experienced 6 times for each fly. n=174-210. **c, Increased seizure prevalence in flies with *Oatp30B* knock down**. The *UAS*-*Oatp30B*^*IR*^ (GD12775) transgenic or the control *UAS-GFP*^*IR*^ were driven with *nSyb-Gal4* and *Ubi-Gal80ts*. Percent +/-SE **** p<0.0001, Log-rank (Mantel-Cox) test, χ^2^ 24.68 for 1 df, n=34-36. **d, Increased post-seizure recovery time in flies with *Oatp30B* knock down**. The *UAS*-*Oatp30B*^*IR*^ (GD12775) transgenic or the control *UAS-GFP*^*IR*^ were driven with *nSyb-Gal4* and *Ubi-Gal80ts*. Mean +/-SEM * p<0.05, Mann Whitney non-parametric test, two tails, n=10-26. Only flies that displayed a seizure within 120 s as in Fig. 4b have been included in the analysis.

The lead associated SNP at the *SLCO5A1* locus has MAF=4.1% and accounts for 10.1% PVE. We next sought to assess whether sub-GWAS significant signals could inform additional contributing genes or pathways and whether there were shared genetic contributions with other psychiatric or epilepsy phenotypes. We selected all variants displaying P ≤ 5 × 10^−4^ and annotated these variants to the transcription start site of the nearest gene using the Ensembl Variant Effect Predictor (v94)^39^, resulting in 855 unique genes; these were used as input both in a gene ontology (GO) enrichment analysis^40-42^ and in FUMA’s GENE2FUNC tool^43^ (v1.3.7). Gene enrichment analysis using AmiGO^41^ identified a 9.3-fold enrichment of associated genes from the presynaptic assembly and organisation gene set (nine out of 23 genes; p=0.03; GO:0099054). These genes were *NRXN1, NLGN1, NLGN4X, PTPRD, FZD1, PCLO, CNTN5, IL1RAPL1*, and *PTEN* (Table 2). The combined PVE for the lead variants annotated to these nine genes is 27.4% (34.1% with the addition of rs73293634 from the *SLCO5A1* locus and rs75042057 from the *PARD3* locus).

**Table 2:** List of top variants annotated to the nine presynaptic assembly genes enriched in the European GWAS of BIS in JME (n=324). *PTPRD*, Protein Tyrosine Phosphatase Receptor Type D; *FZD1*, Frizzled Class Receptor 1; *NLGN1*, Neuroligin 1; *NLGN4X*, Neuroligin 4 X-Linked; *PCLO*, Piccolo Presynaptic Cytomatrix Protein; *CNTN5*, Contactin 5; *NRXN1*, Neurexin 1; *IL1RAPL1*, Interleukin 1 Receptor Accessory Protein Like 1; *PTEN*, Phosphatase and Tensin Homolog.

| Gene | Rsid | Beta | P-value | PVE |
| --- | --- | --- | --- | --- |
| *PTPRD* | rs1781264 | 1.827 | 1.19E-04 | 0.042 |
| *FZD1* | rs916716 | 1.428 | 8.16E-05 | 0.051 |
| *NLGN1* | rs73177088 | 6.191 | 9.95E-04 | 0.044 |
| *NLGN4X* | rs146813567 | -2.898 | 3.06E-04 | 0.039 |
| *PCLO* | rs4728525 | -2.311 | 7.35E-04 | 0.052 |
| *CNTN5* | rs150418619 | 5.806 | 1.06E-04 | 0.051 |
| *NRXN1* | rs1040239 | -1.573 | 1.21E-04 | 0.045 |
| *IL1RAPL1* | rs5943492 | 1.039 | 8.73E-04 | 0.043 |
| *PTEN* | rs112050451 | 5.158 | 1.27E-03 | 0.041 |

Investigation of these 855 genes revealed further specificity regarding localisation and function. First, a comparison with GTEx v8 54 tissue types revealed an enrichment of these genes in the cerebral cortex (-log_10_P>5). Second, we found significant enrichment of these 855 genes with two gene pathways in the Molecular Signatures Database (MSigDB v7.5.1)^44^ identified as being active in murine whole brain^45^ and neuronal progenitor cells^46^. Third, there was significant overlap with genes reported in the GWAS Catalog that contribute to phenotypes relevant to the predominance of JME seizures on awakening, impulsivity and metabolism: chronotype (68 out of 549 genes overlap, p=1.14 × 10^−12^), obesity-related traits (77 out of 706 overlap, p=7.33 × 10^−12^), general risk tolerance (33/247 overlap, p=7.56 × 10^−7^), and adventurousness (23/141, p= 3.15 × 10^−6^). Last, given impulsivity is a major component of ADHD, bipolar disorder and epilepsy, we tested and found that a higher ADHD polygenic risk score (PRS) was significantly associated with a higher BIS-Brief score (p=1.60 × 10^−3^). PRSs for bipolar disorder, generalized and focal epilepsy did not show significant evidence for association with BIS-Brief score (p=0.08, 0.33 and 0.96, respectively). Interestingly, genes nearest ADHD PRS SNPs were not enriched for synaptic assembly genes. Altogether this suggests that the impulsive trait seen in JME is an endophenotype that shares genetic architecture with impulsivity in the general population as well as with individuals diagnosed with ADHD.

This is the first GWAS of trait impulsivity in a neuropsychiatric disorder and we present evidence for the role of SLC05A1 in impulsivity and seizure susceptibility through triangulation^47^ with GWAS, colocalization with gene expression and functional evaluation using an established model of seizure susceptibility in *Drosophila*^48^. There has only been one GWAS of impulsive traits in the general population, which identified genome-wide significant association with variants in the *CADM2* gene, encoding a cell adhesion protein from the SynCam Immunoglobulin superfamily of recognition molecules, important for synaptic organisation and specificity; and association of variants at the *CACNA1I* locus, which has been observed in previous studies with schizophrenia^9^. Our GWAS did not show significant association with the previously reported associated variants at the *CADM2* and *CACNA1I* loci^9^, (p=0.152 for rs139528938 and p=0.32 for rs4522708 which is a SNP with r^2^=0.87 with the reported SNP, rs199694726, in our BIS dataset). Genome-wide summary statistics were not made available to make additional comparisons including assessing polygenic overlap with risk scores.

Previous expression studies show that *SLCO5A1* upregulates gene sets implicated in cell adhesion, synapse assembly and organisation, principally belonging to the cadherin superfamily^7^; and the enrichment for presynaptic assembly and organisation pathway in our dataset includes genes encoding trans-synaptically interacting proteins such as *NRXN1-NLGN1* and *PTPRD*-*IL1RAP1L* that are implicated in a wide range of neuropsychiatric disorders^49,50^. Genetic correlation between ADHD and the BIS-Brief score suggests converging genetic influences across ADHD and epilepsy. Taken together, these results support an important role for specific cell recognition molecules in the organisation of synaptic connections that have an influence in early adulthood as a mechanism for variation in impulsivity across health and disease^51^.

While prefrontal-striatal inhibitory control networks are implicated in impulse control, specifically between mPFC and nucleus accumbens^18,20^, a role for these limbic networks has only been hinted at in epilepsy. Striato-nigral circuits, preferentially involving the ventral striatum, have long ago been implicated in the *regulation* of generalised seizures in rodent models of generalised epilepsy^19^.

Recently, an *initiating* role for cortico-striatal networks in absence seizures with generalized spike-and-wave discharges has been shown in the mouse model of the genetic epilepsy caused by haploinsufficiency of *STXBP1*^52^, specifically by reduced cortical excitatory transmission onto striatal fast spiking interneurons. The impulsivity-like startling and the seizure-like phenotype of the *SLCO5A1/Oatp30B* knockdown in *Drosophila* suggests the genetic co-causality of impulsivity and seizures. This supports the idea that excitatory-inhibitory imbalance in the prefrontal-striatal network may predispose simultaneously to epilepsy and impulsivity substrates and invites new approaches to neuromodulation of generalised seizures.

## Methods

### Human Participants

We collected clinical and genetic data from the Biology of Juvenile Myoclonic Epilepsy (BIOJUME) consortium study (n=864)^25^. We obtained informed consent from all participants and ethical approval from UK Health Research Authority: South Central Oxford C Research Ethics Committee (16/SC/0266) and all other collaborating sites. The SickKids Research Ethics Committee of The Hospital for Sick Children (1000033784) gave ethical approval for this work.

### Barratt Impulsivity Scale -Brief

We collected self-rating of trait impulsivity through the BIS-brief^3,30^, an eight-item scale generating a total score ranging from 8 to 32.

### Genotyping quality control

We genotyped participants’ DNA in four batches (n=702) using the Illumina Omni 2.5 array and performed quality control (QC) using PLINK v1.90b6.18^53^ and custom in-house scripts. Briefly, we removed individuals and variants with call rates below 90%; samples with sex-gender mismatches and/or high heterozygosity; males with heterozygous calls for X chromosome markers (non-pseudoautosomal region); and females with non-missing calls for markers on the Y chromosome. We retained heterozygous calls for mitochondrial markers in both sexes (i.e. due to heteroplasmy). We obtained an unrelated sample by using KING v.2.2.4 software’s^54^ --unrelated option (that is, those with estimated kinship coefficient less than 0.088). We corrected and updated the ped file with all found relationships, and identified markers with Mendelian errors using PEDSTATS 0.6.12^55^. We flagged 399 markers but did not remove those out of Hardy-Weinberg Equilibrium (p < 10^−4^). We conducted principal component analysis adjusted using the kinship matrix output by KING using PC-AiR in the GENESIS v2.16.0 package^56^.

We performed quality control on each genotyping batch separately, followed by removal of ambiguous A/T, G/C SNPs, chr0 SNPs, indels, monomorphic variants, and duplicate variants; and performed strand alignment using Will Rayner’s alignment files (https://www.well.ox.ac.uk/~wrayner/strand/), then merged all batches. We re-analysed and removed cryptic relationships across batches. The final merged set contained 1,489,917 variants (we removed a large number of monomorphic variants), 695 individuals (241 males, 454 females) including 23 related pairs (for association analyses however, an unrelated set is selected).

### Genotype imputation

We used the McCarthy Tools v4.3.0 to prepare the genotype data for imputation (https://www.well.ox.ac.uk/~wrayner/tools/HRC-1000G-check-bim-v4.3.0.zip) using TOPMED as the reference panel (r2@1.0.0) on the TOPMED imputation server^57-59^. We converted coordinates from hg37 to hg38 coordinates using strand files (https://www.well.ox.ac.uk/~wrayner/strand/InfiniumOmni2-5-8v1-4_A1-b38-strand.zip). We merged the pseudoautosomal region (PAR) using PLINK’s --merge-x option and checked variants using the HRC checking tool. We removed a total of 282,660 variants due to no matches in the reference (but still analyzed for association with BIS afterwards), and 1,739,329 variants remained for imputation on the server. We used Eagle v2.4 for phasing, and minimac v4 v1.0.2 for imputation. We kept variants with imputation quality score r^2^>0.4 and minor allele frequency (MAF) > 1% for analysis. A total of 8,950,360 variants remained for association analysis.

### Genome-wide association analysis

We included for analysis 381 individuals who passed phenotype QC with complete BIS-Brief rating. From these, four failed genotyping QC, and one individual was removed due to cryptic relatedness (n=376). The mega-GWAS analysis consisted of a total of 372 unrelated individuals adjusted for sex, genotyping batch, and population stratification (Supplementary Fig. 2). The mega-GWAS was used for colocalization analysis of the genome-wide association peak on chromosome 8. We identified 329 patients as European ancestry (defined as within 6 standard deviations from the 1000 Genomes^60^ European cluster in a principal component analysis). Among these, four patients were missing information on seizure frequency, so we used 324 individuals for the genome-wide association analysis. We adjusted for sex, genotyping batch, age at consent, population stratification, and the frequency of myoclonus or absence seizures. All analyses were conducted on the European GWAS unless noted otherwise. Chromosome X (non-pseudoautosomal region) was analysed with males coded as zero for the reference allele and two for the alternate allele.

### Gene enrichment analysis

Variants with p ≤ 5 × 10^−4^ were annotated to the gene with the nearest transcription start site using the Ensembl Variant Effect Predictor (v94)^39^, which mapped to 855 unique genes. This gene set was used as input both in a gene ontology (GO) enrichment analysis^40-42^ and in FUMA’s GENE2FUNC tool^43^ (v1.3.7), to test for enrichment in annotated pathways. The GO enrichment analysis revealed enrichment of nine presynaptic assembly genes in our dataset, which were followed up with further analyses.

### Phenome-wide association study (PheWAS) analysis

We queried the top associated genome-wide variant and the top associated variant for each of the nine presynaptic assembly enriched genes across PheWAS databases: GWAS Atlas (https://atlas.ctglab.nl/), Global Biobank Engine^61^, PheWeb^62^, and Gene Atlas^63^.

We used PheWeb portals:

- UK Biobank: https://pheweb.org/MGI-freeze2/
- Oxford Brain Imaging Genetics (BIG) Project: http://big.stats.ox.ac.uk/
- fastGWA: https://yanglab.westlake.edu.cn/resources/ukb_fastgwa/imp/
- https://pheweb.org/UKB-SAIGE/

### Polygenic risk score (PRS) analysis

Clumping and thresholding was used to calculate attention deficit hyperactivity disorder (ADHD), bipolar disorder, generalized epilepsy and focal epilepsy PRS in individuals of European ancestry using PLINK v1.9^53^. Four PRS were considered and calculated, corresponding to a Bonferroni-corrected p-value for significance of 0.05/4=0.0125. The source of summary statistics used, variant filtering, clumping and thresholding details are summarized in Extended Data Table 2. PRS values were generated by weighting selected SNPs after clumping and thresholding by the additive scale effect (log^10^ OR), and then summing over the variants. The PRS values were then centred to the mean. Association of PRSs with BIS-Brief was tested using linear regression with age, sex and frequency of absence/myoclonic seizure as covariates in the model.

### Colocalization analysis

We used the Simple Sum 2^34^ and COLOC2^64^ colocalization methods as implemented in LocusFocus^65^ (v1.4.9) to test for colocalization of the genome-wide peaks with eQTL analyses in brain tissues in GTEx v8^12^, PsychENCODE^32^, and fetal brains^33^. For the genome-wide associated locus on chromosome 8, we performed colocalization analysis on both the mega-GWAS and Europeans-only GWAS (Supplementary Fig. 2). The required significance threshold, after multiple testing of all colocalization datasets analyzed was 0.01.

### Domain architecture of SLCO5A1

A BLAST search against the entire Protein Data Base (PDB) identified only one hit with a convincingly high E-value (1e-55) that pointed to the Chain L of the Kazal-like domain containing mice protein (7EEB). The search had a 26% identity and a coverage of 74%. After this hit, the other four identified sequences had E-values > 0.002, clearly distinguishing between significant and non-significant hits. 7EEB is a large complex containing several subunits, among which is *SLCO6C1*, which is the region scoring for *SLCO5A1*.

### Phenotypic variance explained

To assess the phenotypic variance explained (PVE) by a SNP or a group of SNPs, we calculated the partial r^2^ as the proportion of the residual sum of squares (RSS) reduced when adding the SNP (or group of SNPs) to the base regression model with all covariates.

### siRNA probe design and knockdown of *Oatp30B* in *Drosophila melanogaster*

#### Drosophila

Flies were maintained and crossed at 18°C. All ageing was done in a controlled environment of 29°C and 60% humidity.

#### Stocks

*ubiGal80*^*ts*^ // *UAS*-*Oatp30B*^*IR*^ (GD12775) obtained from the VDRC // *w*^*1118*^, *nSybGal4, TubGal4* and *UAS-GFP*^*IR*^ obtained from the BDSC.

#### Lifespan

Lifespan analysis was performed as previously reported^38^. All crosses were maintained at 18°C during the developmental stages of the progeny. Newly eclosed adult flies were collected within 5 days at 18°C. Females and males were pooled together and equally distributed within vials.

#### Motor behaviour assay

Single fly tracking was carried out as previously described^38^. In each of 3 experiments, up to 12 flies per genotype, aged 15 days (adult stage) at 29°C to allow RNAi expression and knock-down, were placed into individual round 6-wells arenas. The protocol used consisted of 6 stimuli events equally split during a period of 2 h and 15 min, the first one starting after 30 min of recording and the last one 30 min before the end of the protocol. Each stimuli event was composed of 5 vibrations of 200 ms spaced by 500 ms. The x/y position of each single fly was tracked and analysed using the DART software in order to evaluate the relative speed and activity before, during and after the stimuli event. The speed analysis was used for the “Stimuli Response Trace” and the general activity used to deduce “Active Speed”, “Mean Bout Length” and “Inter-Bout Interval”, using a custom-made modification of the DART software^37^.

#### Heat-induced seizure assay

Flies aged 15 days at 29°C to allow RNAi expression and knock-down were isolated into new plastic vials without food for 10-20 min before immersion in a 42°C water bath for 120 seconds. Each tube was video recorded during and post immersion and seizures were defined as a period of brief leg twitches, convulsions and failure to maintain standing posture. Flies were, thereafter, allowed to recover at room temperature and the time to recover from seizure was calculated only for flies that had undergone seizures. All experiments were randomised and double-blinded.

#### RNA extraction and qPCR

RNA was extracted as previously reported^66^ from 15 adult flies of both sexes, aged 15 days at 29°C to allow RNAi expression and knock-down, using TriZol (Thermo-Fischer). cDNA was generated using SuperScript III Reverse Transcriptase (Thermo-Fischer). Quantitative PCR was performed in combination with qPCRBIO SyGreen Blue mix (PCR Biosystems) on Quantstudio 7 from real-time PCR system (Thermo-Fischer). *eIF4a* was used as housekeeping control. The following oligos were used: *Oatp30B* Fw (GAATCCGACCAACCGCCTGA), *Oatp30B* Rv (ATGGATTCCTGCCGCCTGTG), *eIF4a* Fw (CGTGAAGCAGGAGAACTGG), *eIF4a* Rv (CATCTCCTGGGTCAGTTG).

## Data Availability

eQTL data are available for download for GTEx (https://gtexportal.org/home), PsychENCODE (http://resource.psychencode.org/), and fetal brains (https://doi.org/10.6084/m9.figshare.6881825). GWAS summary statistics for this study are available for download from our website (https://lab.research.sickkids.ca/strug/softwareandresources/).

http://resource.psychencode.org/

https://doi.org/10.6084/m9.figshare.6881825

https://lab.research.sickkids.ca/strug/softwareandresources

## Author Contributions

LJS and DKP contributed to conception and study design. NP, DR, AS, AC, AH, LJS and DKP contributed to data management and project administration. DA, JPC, CPB, CYF, JG, DAG, CD, FM, KH, KSL, JK, AO, MR, KKS, GR, MS, IT, RT, JZ, MPR, LJS and DKP contributed to acquisition of study data. NP, DR, AS, MF, LJS, and DKP contributed to analysis of data. NP, DR, AS, MF, LJS, and DKP contributed to drafting the manuscript. Members of the BIOJUME consortium are listed in the appendix.

## Acknowledgements

This work was supported by the Canadian Institutes of Health Research: Biology of Juvenile Myoclonic Epilepsy 201503MOP-342469 (DKP, LJS) and 201809FDN-407295 (LJS); UK Medical Research Council, Centre for Neurodevelopmental Disorders MR/N026063/1 (DKP, MPR); UK Medical Research Council, Programme Grant MR/K013998/1, (MPR); PhD stipend from UK Medical Research Council and the Sackler Institute for Translational Neurodevelopment (AS); NIHR Specialist Biomedical Research Centre for Mental Health of South London and Maudsley National Health Service Foundation Trust (DKP, MPR); UK Engineering and Physical Sciences Research Council, Centre for Predictive Modelling in Healthcare (EP/N014391/1 (MPR)); DINOGMI Department of Excellence of MIUR 2018– 2022 (legge 232 del 2016 (PS)); Wales BRAIN Unit and Research Delivery Staff funded by Welsh Government through Health and Care Research Wales (KH); Biomarin srl, ENECTA srl, GW Pharmaceuticals, Kolfarma srl. and Eisai (PS); South-Eastern Regional Health Authority, Norway (Project Number 2016129 (JK)); The Research Council of Norway (Project Number 299266 (MS)); Epilepsy Research UK (RT, MR); Health & Care Research Wales (MR), Wales Gene Park (MR), Abertawe Bro Morgannwg University NHS R&D (MR); UCB (GR); Nationwide Children’s Hospital (DAG); Odense University Hospital (JG); University of Southern Denmark (17/18517 (CPB)); Grants NC/V001051/1 from the NC3Rs (MF), European Union’s Horizon 2020 Research and Innovation Programme (765912 - DRIVE - H2020-MSCA-ITN-2017 (HJ)) and Action Medical Research (GN2446 (HJ, MF)).

## Extended Materials: Figures

**Extended Data Fig. 1:**
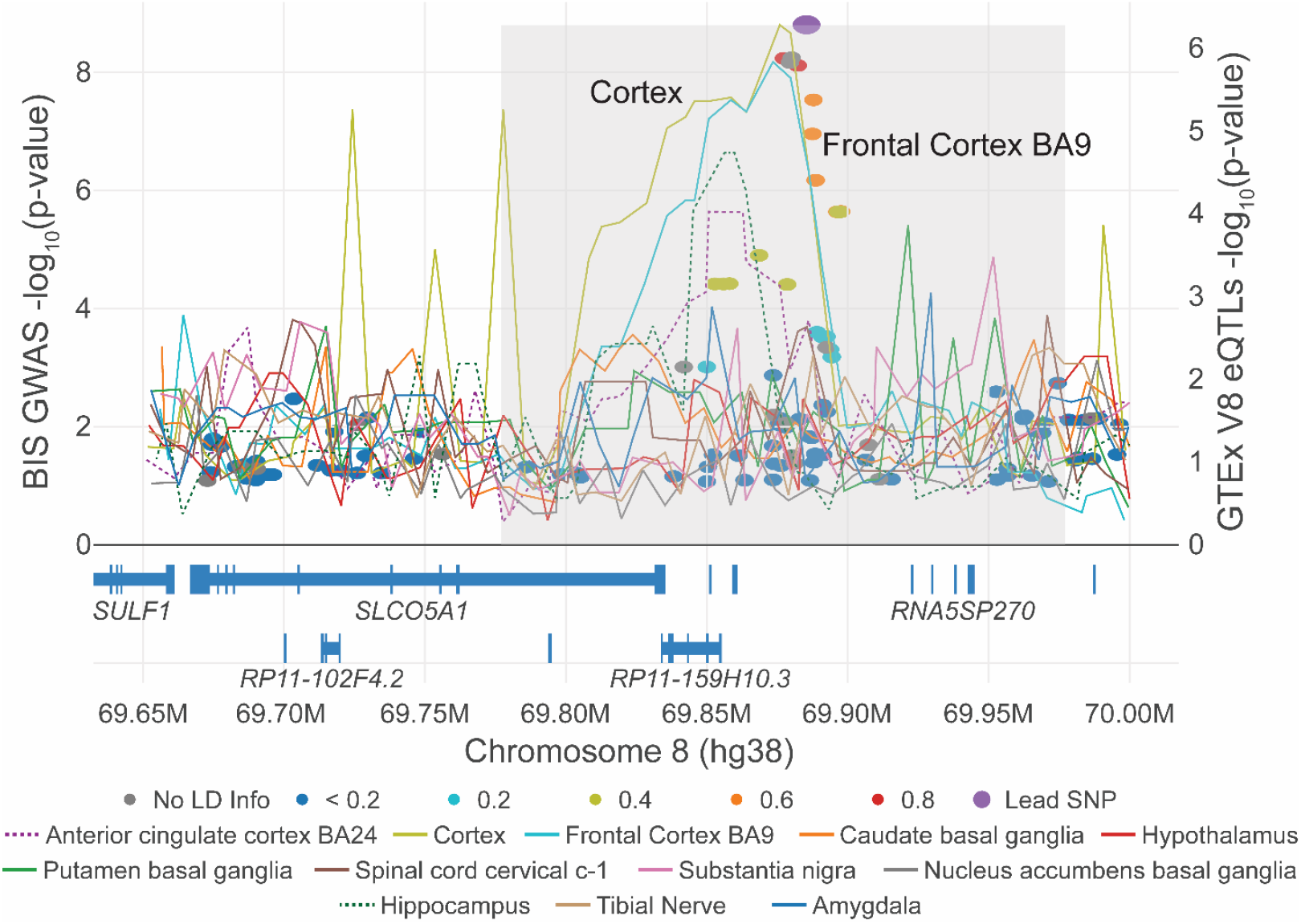
Colocalization figure from LocusFocus^65^ for the *SLCO5A1* gene. Same as in main Figure 3A, but circles depict the GWAS with BIS in the European subset (n=324). Colocalization analysis results reveal colocalization with GTEx^12^ V8 brain cortex (-log_10_ Simple Sum P-value (SSP) = 1.36), although this colocalization does not pass the multiple testing significance threshold of -log_10_SSP=2.0 for testing colocalization across eQTLs from GTEx^12^, PsychENCODE^32^ and fetal brain eQTLs from ^33^.

**Extended Data Fig. 2:**
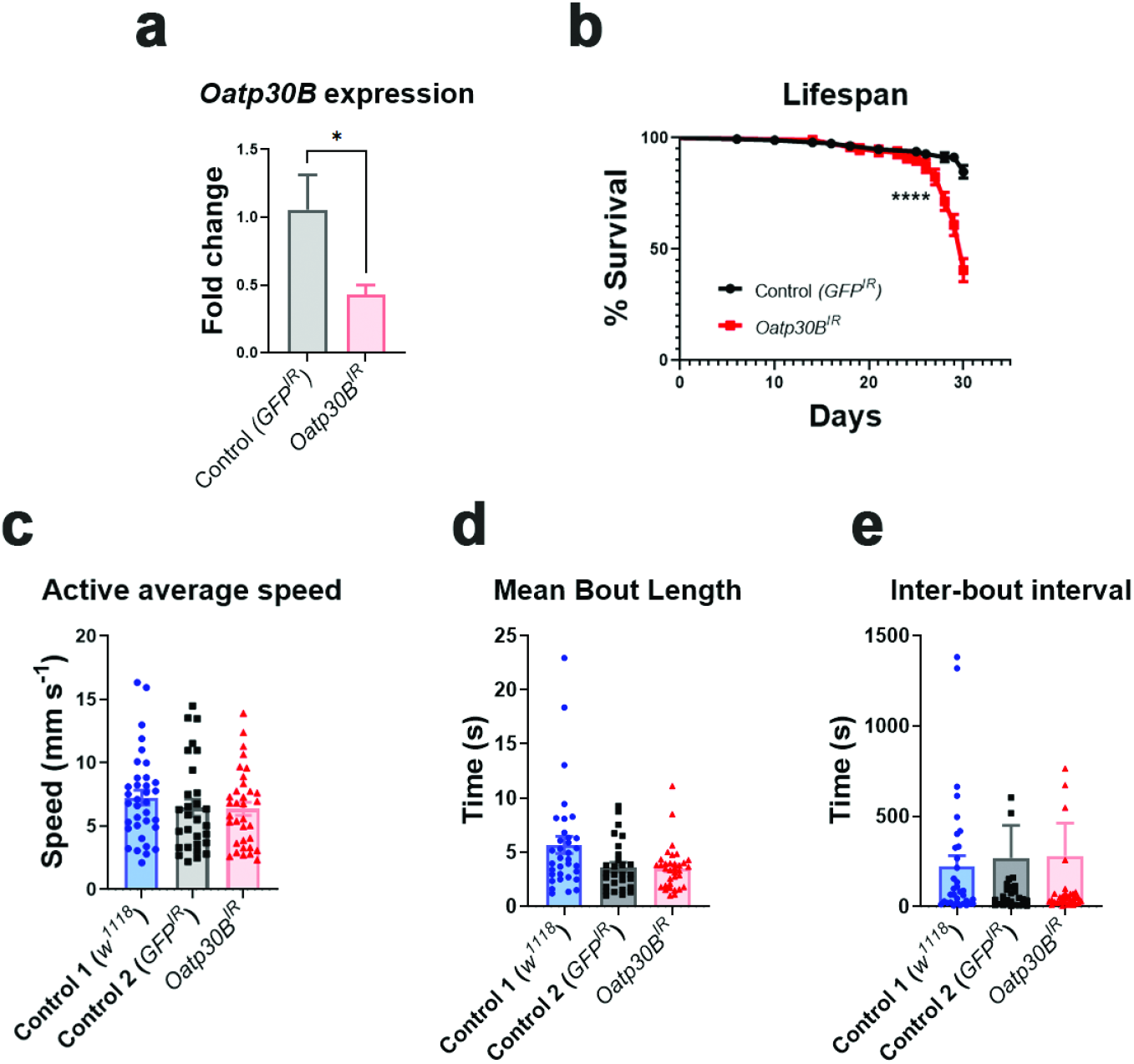
a, Oatp30B mRNA levels as measured by qPCR. The *UAS*-*Oatp30B*^*IR*^ (GD12775) transgenic or the control *UAS-GFP*^*IR*^ were driven with *Tub-Gal4* and *Ubi-Gal80ts*. The graph reports data from 3 biological samples (n=10 flies, both males and females) and 5 technical replicates. Mean +/- SEM * P<0.05, Unpaired t-test, one-tailed. **b, Reduced lifespan in flies with *Oatp30B* knock-down**. The *UAS*-*Oatp30B*^*IR*^ (GD12775) transgenic or the control *UAS-GFP*^*IR*^ were driven with *nSyb-Gal4* and *Ubi-Gal80ts*. Percent +/-SE **** P<0.0001, Log-rank (Mantel-Cox) test, χ^2^51.74 for 1 df, n=119-194. **c, Unchanged speed during action**. The *UAS*-*Oatp30B*^*IR*^ (GD12775) transgenic or the control *UAS-GFP*^*IR*^ were driven with *nSyb-Gal4* and *Ubi-Gal80ts*. The *w*^*1118*^ strain is a control for the genetic background in absence of transgenes. **d, Unaffected duration of single action bouts**. Flies as in c. **e, Unaffected rest interval in between single action bouts**. Flies as in c.

## Extended Materials: Tables

**Extended Data Table 1:**
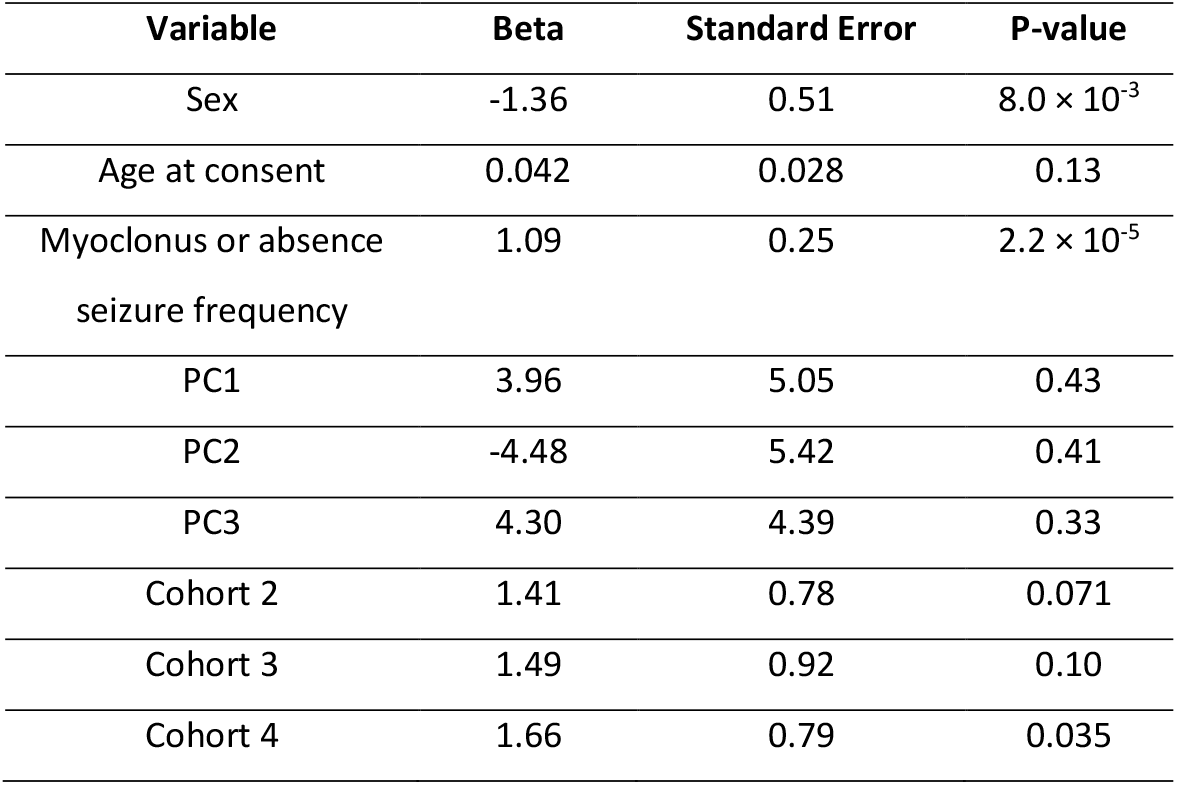
Univariate analysis with BIS. A total of 324 European individuals with JME were tested for association for each of the variables shown with BIS. We have previously shown the association of sex and seizure frequency with BIS (ref). Seizure frequency was used as a marker of controlled seizures and was defined as missing if there was no reported myoclonus frequency; otherwise, it was the maximum observed frequency for myoclonus or absence seizure as follows: daily seizures=3; weekly=2; less than weekly=1; none (currently or ever)=0. Cohort refers to the genotyping batch; samples recruited were genotyped in four batches at different time points.

**Extended Data Table 2:** Methods applied for the computation of PRS.

| Disease | Study | Summary statistics |  | Variant filtering | BIOJUME<br>Variant<br>Filtering | Clumping + Thresholding |  |
| --- | --- | --- | --- | --- | --- | --- | --- |
|  |  | Sample size | Ethnicity |  |  | Clumping | p-value<br>Threshold |
| ADHD | Demontis et al., 2019<sup>70</sup> | 19,099 cases & 34,194 controls | European | MAF $\leq 0.05$<br>$r^2 \leq 0.8$<br>Indels<br>Ambiguous SNPs<br>Multi-allelic SNPs<br>MHC region | $r^2 \leq 0.5$ | $r^2$: 0.3<br>radius: 500 kb | 0.1 |
| Bipolar Disorder | Stahl et al., 2019<sup>71</sup> | 20,352 cases and 31,358 controls | European | MAF $\leq 0.05$<br>Info $\leq 0.9$<br>Indels<br>Ambiguous SNPs<br>Multi-allelic SNPs<br>MHC region except for the<br>most significant associated<br>SNP (rs36034627,<br>Chr6:27,269,584, T>G) | $r^2 \leq 0.5$ | $r^2$: 0.1<br>radius: 500 kb | 0.2 |
| Generalized epilepsy* | ILAE Consortium, 2018<sup>72</sup> | 15,212 cases & 29,677 controls | Majority<br>European | MAF $\leq 0.02$<br>$r^2 \leq 0.3$<br>Ambiguous SNPs | $r^2 \leq 0.5$ | $r^2$: 0.1<br>radius: 500 kb | 0.5 |
| Focal epilepsy* | ILAE Consortium, 2018<sup>72</sup> | 15,212 cases & 29,677 controls | Majority<br>European | MAF $\leq 0.02$<br>$r^2 \leq 0.3$<br>Ambiguous SNPs | $r^2 \leq 0.5$ | $r^2$: 0.1<br>radius: 500 kb | 0.5 |

MHC region: The major histocompatibility complex region, chr6:25-34Mbp
ILAE: International League Against Epilepsy
The methods applied to calculated generalized and focal epilepsy PRS are based on Leu et al. 2019<sup>73</sup>.

## Appendix: BIOJUME Consortium

Sites and site investigators included in the BIOJUME consortium.

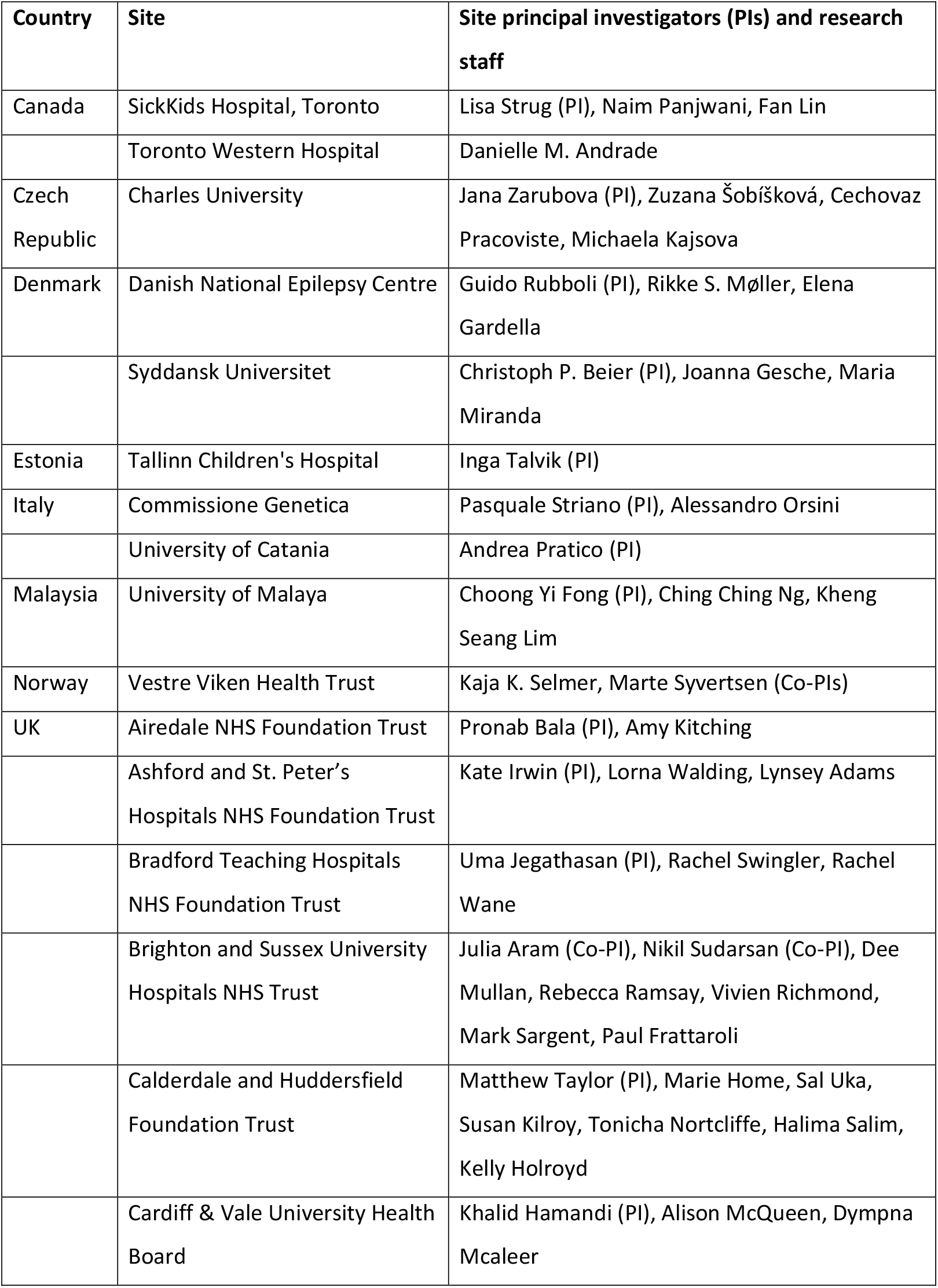
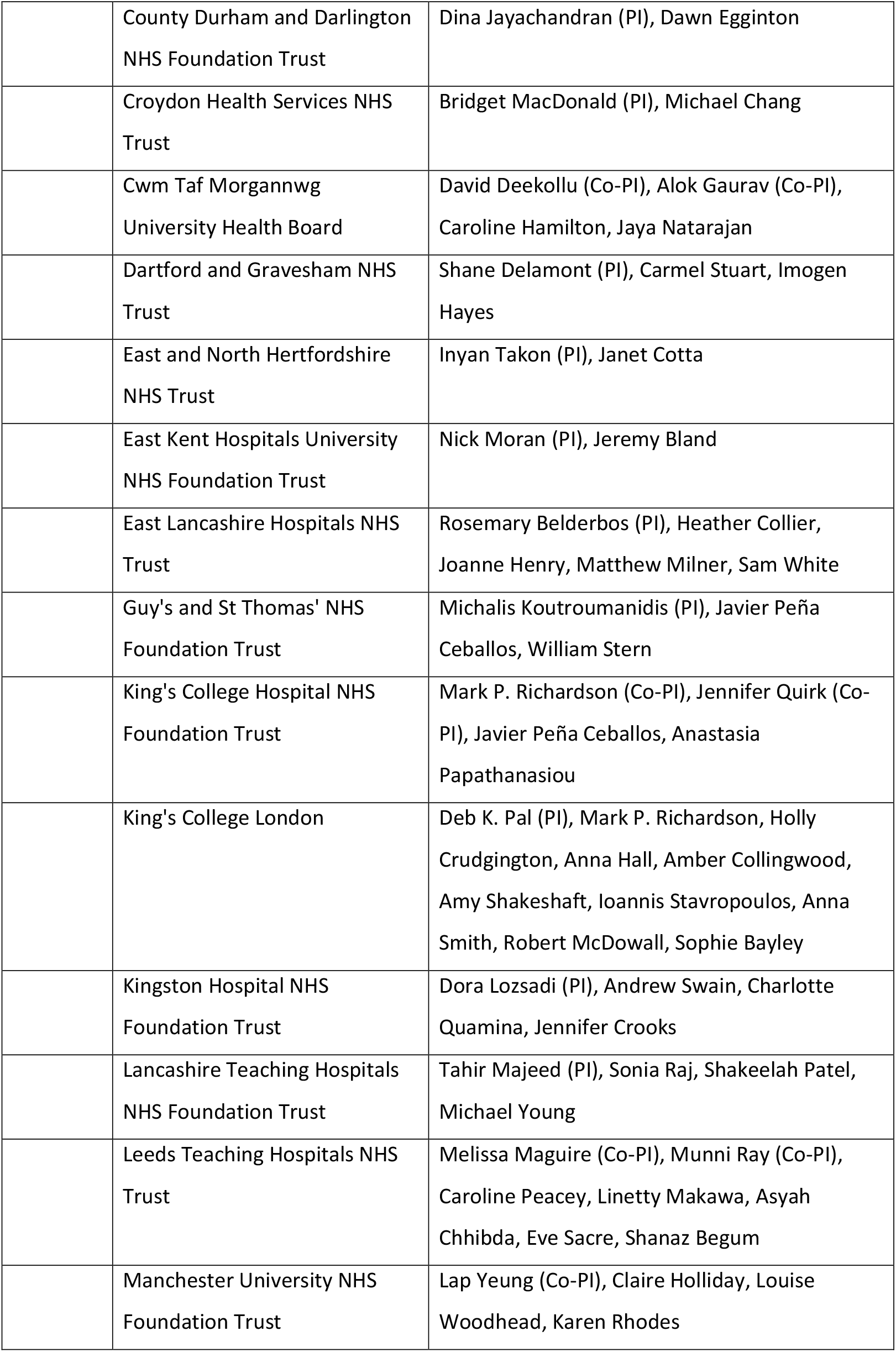
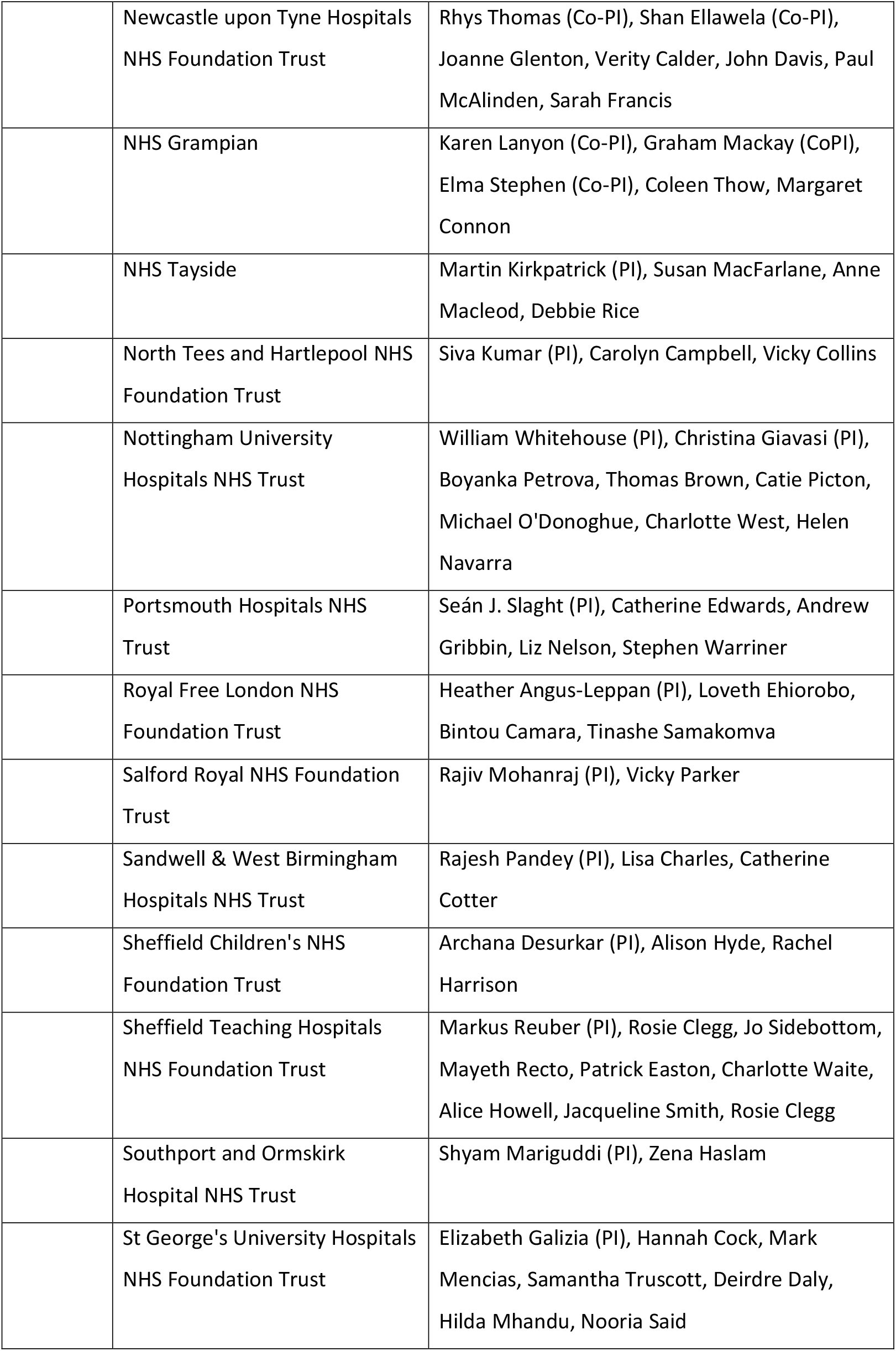
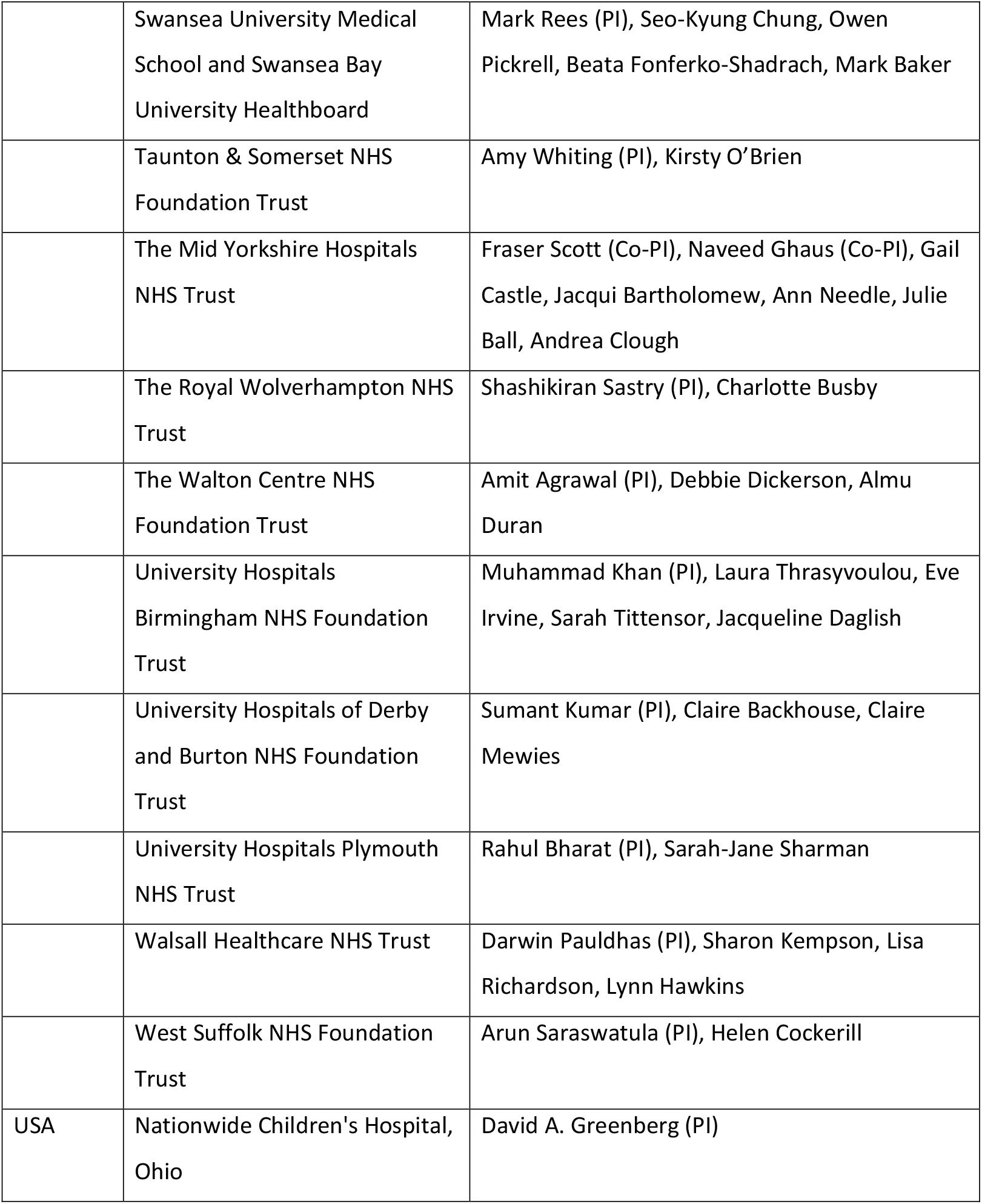

