## Supplementary Fig. 1 for "*SLCO5A1* and synaptic assembly genes contribute to impulsivity in juvenile myoclonic epilepsy"

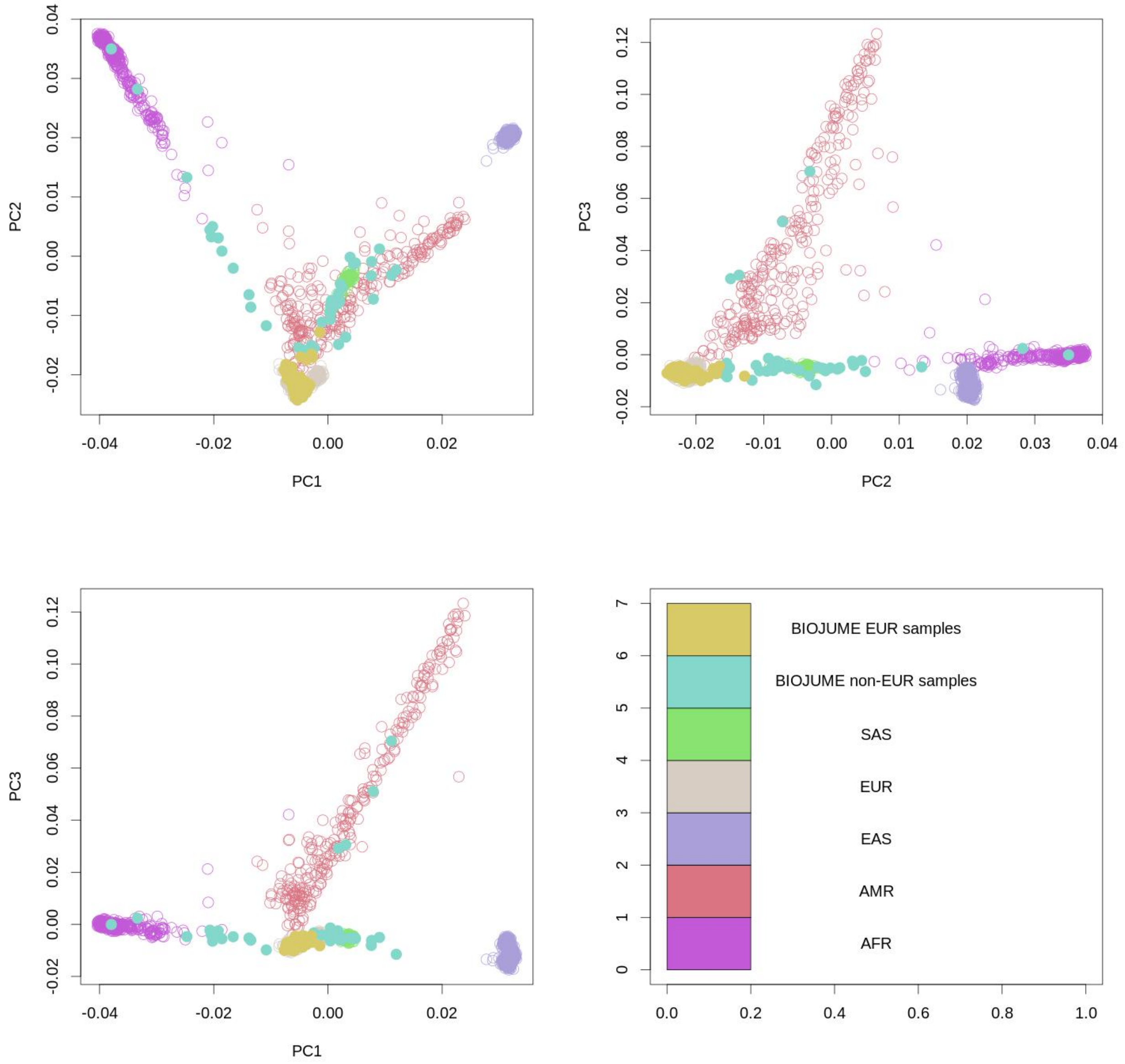


**Supplementary Fig. 1. Principal component analysis (PCA) of all study samples with the 1000 Genomes (phase 3).** BIOJUME samples who were admixed or AMR were kept if they were within 6 standard deviations from the European cluster. SAS, South Asian; EUR, European; EAS, East Asian; AFR, African; AMR, Ad Mixed American.
