## Supplementary Fig. 2 for "*SLCO5A1* and synaptic assembly genes contribute to impulsivity in juvenile myoclonic epilepsy"

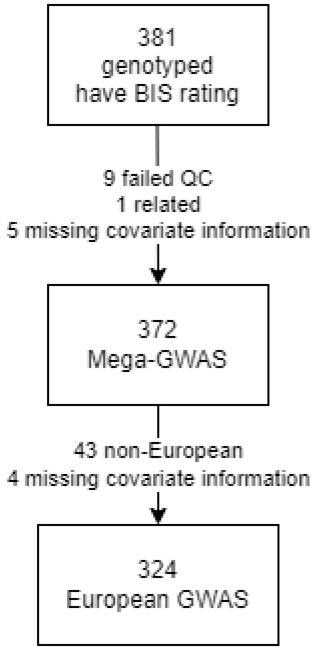


**Supplementary Fig. 2. Flowchart of inclusion for the mega-GWAS and the European GWAS.** The mega-GWAS includes individuals of all ethnic backgrounds, while the main European GWAS includes those individuals within six standard deviations from the 1000 Genomes-defined European cluster.
